# Protocol for the Houston Hospital-Based Violence Intervention Program

**DOI:** 10.1101/2025.05.18.25327870

**Authors:** Alexander Testa, Eresha Bluth, Latanya Monroe, Karlton Harris, Sarah Beth Abbott, Mary E. Aitken, Erin E. Fox, Brian Heckler, Lillian S. Kao, Ruosha Li, Susannah (Meg) Michael, Heidi M. McPherson, Marisol Nieves, Christian P. Owen, Kevin Rix, Vanessa Schick, Shreela V. Sharma, Carlie Stratemann, Anne Marie V. Thompson, Jack Tsai, Zixi Yang, Sandra McKay

**Affiliations:** Department of Management, Policy and Community Health, School of Public Health, University of Texas Health Science Center at Houston, Houston, TX, United States of America; Department of Pediatrics, McGovern Medical School, University of Texas Health Science Center at Houston, Houston, TX, United States of America; The Forgotten Third, Houston, TX, United States of America; Children’s Memorial Hermann Hospital, Houston, TX, United States of America; Department of Surgery, McGovern Medical School, University of Texas Health Science Center at Houston, Houston, TX, United States of America; Department of Biostatistics and Data Science, School of Public Health, University of Texas Health Science Center at Houston, Houston, TX, United States of America; Memorial Hermann Hospital, Houston, TX, United States of America; Health Equity Collective, School of Public Health, University of Texas Health Science Center at Houston, Houston, TX, United States of America; Department of Research, Cizik School of Nursing, University of Texas Health Science Center at Houston, Houston, TX, United States of America; Department of Health Promotion & Behavioral Sciences, School of Public Health, University of Texas Health Science Center at Houston, Houston, TX, United States of America; Department of Epidemiology, School of Public Health, University of Texas Health Science Center at Houston, Houston, TX, United States of America; Michael & Susan Dell Center for Healthy Living, School of Public Health, University of Texas Health Science Center at Houston, Houston, TX, United States of America; National Center on Homelessness Among Veterans, U.S. Department of Veterans Affairs, Washington, DC, United States of America; Department of Psychiatry, Yale University School of Medicine, New Haven, CT, United States of America

## Abstract

Firearm violence is a leading cause of injury and mortality in the United States. Hospital-based violence intervention programs (HVIPs) are a promising public health strategy designed to reduce recurrent violence by engaging patients during hospitalization and connecting them to support services after discharge. This protocol describes the design and implementation of the Houston Hospital-Based Violence Intervention Program (Houston-HVIP), which will be evaluated by a randomized controlled trial conducted at a Level 1 trauma center in Houston, Texas. The study plans to enroll individuals aged 16 to 35 who present with gunshot wounds (GSW) at the Level 1 trauma center. Participants are randomized to either a treatment group receiving six months of intensive case management with direct referrals to social services or a control group receiving usual care, which involves indirect referral and limited case management. The primary outcome is a composite measure of an individual’s exposure to firearm violence via (a) self-report, (b) hospital admission records, and (c) mortality records.

Secondary outcomes measured at the individual level assess violent reinjury, attitudes toward violence, post-traumatic stress, aggression, and self-rated health. Outcomes are assessed at baseline and 3-, 6-, 9-, and 12-months post-enrollment. The study will enroll 274 participants and include both quantitative and qualitative assessments to evaluate program impact and participant experience. This protocol aims to contribute to the design and implementation of HVIPs in large Level 1 trauma centers.

## Introduction

Firearm violence is a critical public health crisis in the United States (US), resulting in approximately 46,000 deaths in 2023,^1^ as well as tens of thousands of additional individuals who are admitted to the hospital for non-fatal gunshot wounds (GSW) annually.^2,3^ GSWs result in long-term physical, functional, psychological, and social consequences,^4^ as well as an economic cost estimated at approximately $500 billion annually,^5^ with the median medical charge for a single GSW estimated at over $53,832.^6^ Accordingly, interventions aimed at reducing firearm violence and hospital admission for GSWs are essential components of a comprehensive public health response.

Hospital-based violence intervention programs (HVIPs) are a novel strategy with the potential to mitigate firearm violence.^7–9^ HVIPs operate within trauma centers and aim to reduce the risk of recurrent violent injury by offering services to violence survivors before and following hospital discharge. These programs integrate case management, social service referrals, and community-based partnerships to support victims during their recovery and address underlying factors that contribute to violence.^10^ Despite the increased prevalence of HVIPs emerging throughout the US in recent years and evidence of their potential benefits for reducing violent reinjury and retaliatory violence,^9,11,12^ rigorous evaluations of their effectiveness in reducing firearm-related harm are still needed, with a particular lack of adequately powered randomized controlled trials (RCTs).^8,13^

In 2023, the University of Texas Health Science Center at Houston (UTHealth Houston) received funding from the National Institutes of Institute of Nursing Research (NINR) via award UG3NR021232 to develop and evaluate the Houston Hospital-Based Violence Intervention Program (Houston-HVIP) at a Level 1 trauma center in Houston, Texas. The Houston-HVIP follows a two-phase research framework, beginning with an initial development and pilot phase to refine the intervention model (2023-2025), and followed by an anticipated RCT (2025-2028) to evaluate its impact on the primary outcome of firearm violence, as well as secondary outcomes of (a) violent reinjury, (b) attitudes toward violence, (c) post-traumatic stress, (d) aggression, and (e) self-rated health. This protocol paper provides an overview of the Houston-HVIP study design, objectives, methodology, and expected outcomes.

## Objectives and Aims

In 2022, the Community Firearm Violence Prevention (CFVP) Network was established with support from the National Institutes of Health (NIH). The CFVP Network comprises a Coordinating Center, a Steering Committee, and multiple research initiatives to evaluate community-based strategies to prevent firearm violence and associated injuries.^14^ These initiatives foster collaboration between academic institutions and community organizations to develop, implement, and assess innovative, community-driven interventions to reduce firearm violence.

Research studies within the CFVP Network follow a structured two-phase research framework. The initial two-year developmental and pilot phase (UG3) focuses on developing, refining, and pilot testing the intervention. This is followed by a three-year (UH3) phase to evaluate the intervention’s effectiveness. Within the CFVP network, three projects were awarded funding from 2022-2027 (Cohort 1); three additional projects were funded from 2023-2028 (Cohort 2).

As part of Cohort 2, UTHealth Houston began the development of the Houston-HVIP in September 2023. The first two years (2023-2025) prioritized coalition-building by engaging internal and external stakeholders to guide program development and oversee its implementation. Additionally, this phase included a pilot study to test and refine the intervention. Upon completion of the pilot phase, a RCT with 274 GSW patients who present to the hospital with a GSW is planned to begin in September 2025. The UH3 phase has two aims:

**Aim 1:** To determine the impact of the Houston-HVIP on the primary outcome of firearm violence.
**Aim 2:** To determine the impact of the Houston-HVIP on the secondary outcomes of (a) violent reinjury, (b) attitudes toward violence, (c) post-traumatic stress, (d) aggression, and (e) self-rated health.

## Setting

Data collection for this study is scheduled for September 2025, with an 18-month recruitment period and a 12-month follow-up period. The study will occur at a large Level 1 trauma center within the Texas Medical Center—the world’s largest medical complex—in Houston, Texas.^15^ The hospital is one of only two adult Level 1 trauma centers serving a population of over 7 million individuals. A review of hospital records found that 2,609 GSW patients were admitted between 2021 and 2024. In 2024, 685 persons presented to the hospital with a GSW, including 435 in our inclusion age range of 16-35. A distribution of GSW in 2024 by age is presented in Appendix A, which shows that 16-35 represents the highest prevalence of GSWs.

## Eligibility Criteria

To be eligible for inclusion in the study, participants must meet the following criteria: (1) present to the hospital with a GSW, (2) be between 16 and 35 years of age, (3) speak English or Spanish, and (4) live within Harris County, Texas (the county which encompasses Houston).

Exclusion criteria include: (1) GSW mechanism is (a) self-inflicted or (b) unintentional gunshot injury (i.e., being shot with a firearm without evidence of intent to harm), (2) being in police custody during the hospital admission, (3) primary residence is outside of Harris County, TX, and (4) being unable to provide informed consent due to inhibited psychological or development state.

## Eligibility Screening

Program staff conducts eligibility screening by reviewing electronic medical records (EMRs) to identify patients presenting to the hospital with GSW. Eligibility screening occurs Monday through Friday (8:00 am – 5:00 pm) for GSW patients that presented to the hospital during the previous 24 hours. On Monday mornings, staff complete additional reviews for cases arriving from Friday evening (after 5:00 pm) to Monday at 8:00 am. The EMR review includes key patient details such as name, sex, race/ethnicity, mechanism of injury, hospital location (if admitted), and contact info if discharged. Following the initial EMR review, program staff consult with the patient care team—including nurses, physicians, residents, and advanced practice providers—to assess eligibility and the patient’s ability to provide informed consent, including factors such as pain levels, sedation status, and overall demeanor, which are critical to determining the patient’s readiness and willingness to learn about and consent to the program. If the medical team indicates that a patient is not yet in a condition to participate, program staff document the patient’s medical status using Research Electronic Data Capture (REDCap)^16,17^ to schedule a follow-up assessment to be completed by program staff the next business day. This process repeats until a patient is deemed medically stable and able to provide consent and approached by a case manager to establish rapport and begin recruitment.

## Recruitment

Study recruitment procedures are detailed in Figure 1 and Figure 2. If a patient meets the initial eligibility criteria screening and no concerns are raised by medical staff regarding their ability to provide informed consent, a case manager will introduce the study at the bedside. Ideally, this introduction occurs during the initial interactions between the case manager and the patient. However, in some cases, multiple conversations may be necessary to establish rapport, build trust, and generate interest in the study. Case managers follow the rapport-building protocol outlined in Appendix B to facilitate this process.

**Figure 1:**
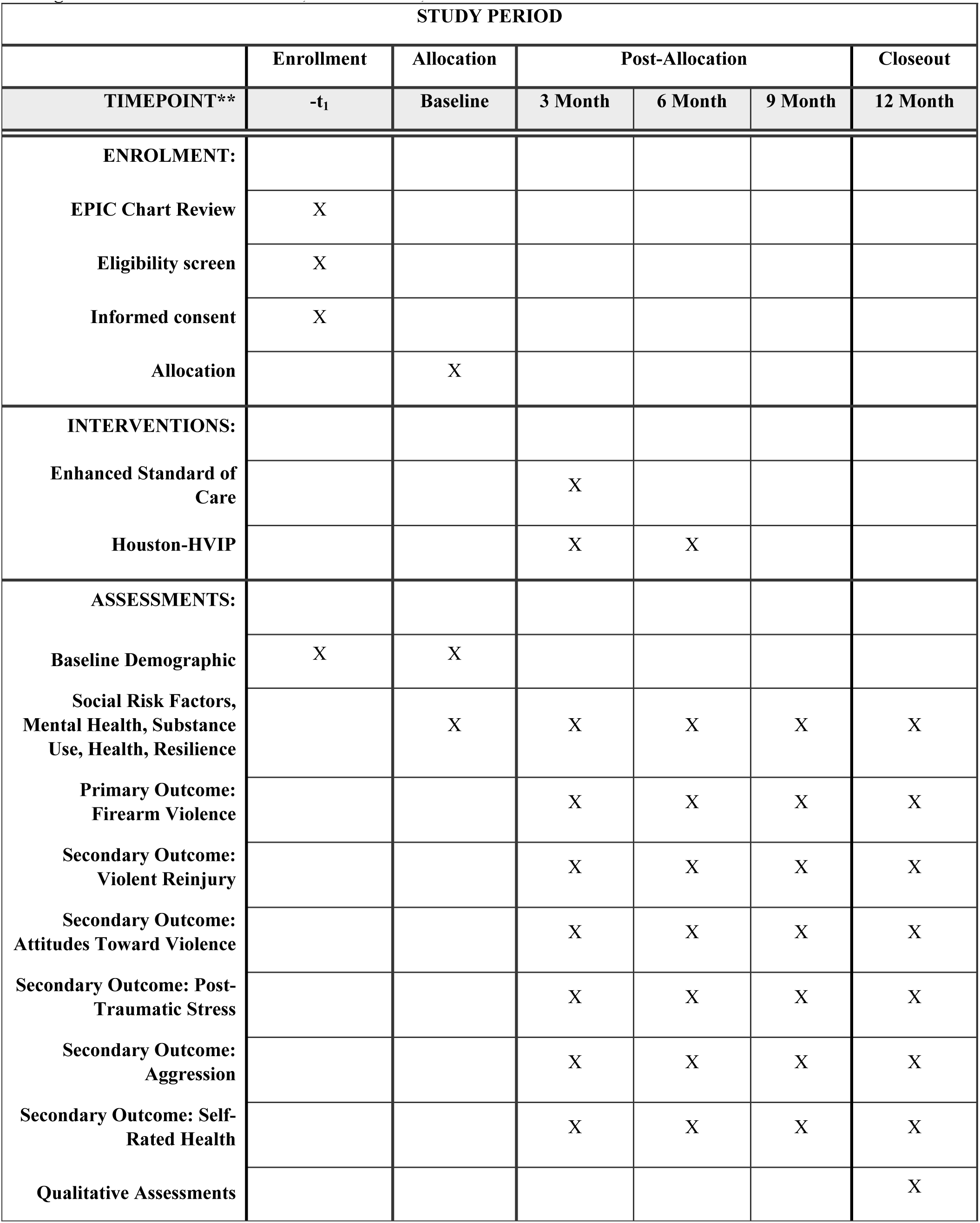
Schedule of enrolment, interventions, and assessments

**Figure 2:**
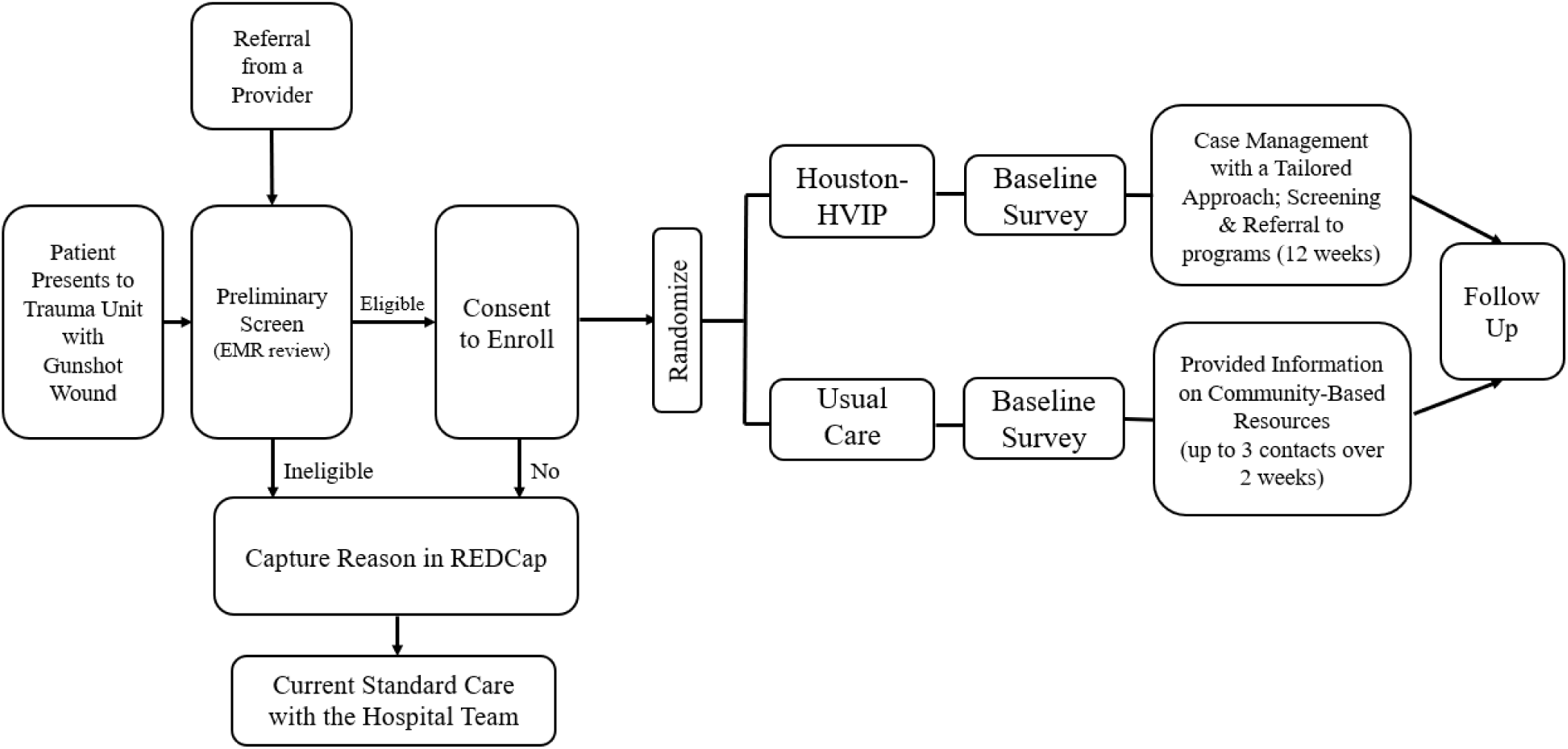
Study Recruitment Procedures

To confirm eligibility, the case manager administers a four-item eligibility screener (see Appendix C). Once eligibility is confirmed, the patient will receive detailed information about the study and their right to decline participation. After reviewing and discussing the informed consent form, the case manager will obtain written consent from interested participants.

Additionally, participants will complete a contact form for follow-up purposes, which includes (a) primary and secondary phone numbers, (b) email addresses, (c) home addresses, (d) secondary residential addresses, (e) emergency contact(s) information, and (f) social media usernames (if applicable).

For patients who meet eligibility criteria but are discharged before engaging with a case manager at the bedside, the case manager will make up to eight follow-up contact attempts over two weeks using the contact information in the EMR. Contact attempts are made via phone calls every other day. If phone calls are not answered, a voice mail is left, and an SMS text message is also sent (if the number listed is a cell phone). The case manager will attempt to reach individuals via telephone at various times and on different days of the week. Patients who decline participation at any time are no longer contacted. In addition, information on study materials and instructions on contacting the case manager are mailed to the participant at the address listed in the EMR. If a discharged patient expresses interest in participating after being contacted, arrangements will be made to meet them in person at the hospital or virtually via Microsoft Teams, Zoom, or a telephone call to discuss study details and obtain informed consent via an electronic consent process. A patient is eligible to enroll for up to one month following discharge.

Additionally, all trauma staff have been educated on the Houston-HVIP project and may share the study information with GSW patients. In these cases, medical staff have been trained and provided a badge with a QR code that links to information about the study and study staff contact information. If any hospital medical staff believes a patient is eligible for the program, a referral can be made by sending a patient’s information to Houston-HVIP study staff to begin the screening and consent process.

## Intervention/Treatment

The RCT will be a parallel-group, two-arm trial with a 1:1 allocation ratio. After informed consent is obtained, participants are randomly assigned to either the treatment condition (Houston-HVIP) or the control condition (Standard of Care). Randomization is conducted in REDCap using permuted block randomization with varying block sizes. The randomization module in REDCap is pre-programmed with a computer-generated sequence to minimize selection bias and maintain allocation concealment. The targeted enrollment for the study is 274 participants (137 enrolled in each arm). The study will employ three full-time case managers, ensuring that each case manager is assigned no more than 10 to 12 high-intensity cases (i.e., cases in the treatment condition) at any given time during the six-month treatment period. Enrolled participants are randomly assigned to a case manager. This caseload aligns with the Health Alliance for Violence Intervention (HAVI) recommendations for the maximum recommended number of high-intensity cases per case manager and the appropriate duration of case management services for HVIPs.^18^ Due to the nature of the planned intervention, participants and case managers will not be blinded to group assignment.

### Control Condition: Enahnced Usual Care

The control condition is a “light-touch” intervention, where participants receive a one-off list of resources.^19^ However, because of ethical concerns about identifying unmet social needs without offering support, participants will be provided with brief follow-up support from a case manager in the form of up to three contact attempts over two weeks to help answer questions about the provided information and address challenges navigating desired services. Accordingly, this goes beyond the typical standard care provided by the hospital system, and thus, we have conceptualized the control condition as “enhanced usual care.” Participants randomized to the control arm will receive the following:

1. **Indirect Resource Referral:** A list of community resources that address social needs (i.e., educational and financial needs and referrals for job training, educational support, housing assistance, and financial assistance programs).
2. **Trauma Survivors’ Network Engagement:** Information on opportunities for long-term engagement with the hospital’s trauma survivors’ network, which delivers peer support and various resources for survivors of traumatic injury, including twice-monthly virtual support groups and an online self-management/recovery course.
3. **Follow-Up Support:** A case manager will conduct up to three follow-up contacts over two weeks post-discharge to offer assistance with navigating community-based services. Contacts are made by a phone call; if a participant does not answer, both a text message and an email are sent.

### Treatment Condition: Houston-HVIP Program

The treatment condition is an intensive “high-touch” intervention where case managers provide ongoing assistance and direct referral to community-based organizations over six months.^19^ The treatment condition is designed to meet the criteria for a high-intensity social needs intervention, which, based on a recent systematic review, includes (a) ≥8 contacts between a case manager and study participant, (b) meetings every 2 weeks, (c) planned meeting duration of ≥30 minutes, and (d) program duration of ≥6 months.^20^ Participants in the Houston HVIP intervention arm will receive the following (see Table 1 for additional details on the week-by-week case management plan):

1. **Case Management and Risk Assessment:** A case manager will meet with each participant to assess social needs and risk factors for violent reinjury. This assessment will inform an individualized case management plan (see Appendices D and E).
2. **Personalized Social Service Linkages:** Case managers will work with a participant to identify their most immediate needs and use a direct referral model (i.e., the case manager forwards a patient’s contact information to a referral site contingent on the patient’s consent) to connect participants with tailored community-based services, addressing key social needs (i.e., job training, educational support, housing assistance, nutritional assistance, financial assistance).^21^ Every four weeks, the case manager will revisit the social service linkage plan and readdress social needs to ensure that the most immediate and pressing needs are consistently addressed. After three months, the case manager will also provide support in navigating to resources addressing the participants’ long-term social needs.
3. **Enhanced Violence Prevention Support:** Based on responses to the risk-assessment screener (see Appendix E), participants identified as being at elevated risk for violent reinjury will be referred to violence interrupter services.
4. **Trauma Survivors’ Network Engagement:** As with the control arm, participants will receive information about long-term engagement opportunities within the hospital’s trauma survivors’ network.

**Table 1:** Week-by-Week Case Management Plan

| **Week** | **Case Management Activities** |
| --- | --- |
| Week 1 | - Review participant screeners and identify immediate social needs<br>- Introduce available HVIP services<br>- Refer to violence interruption services if violence risk assessment score >3<br>- Begin developing an individualized case management plan with the participant |
| Week 2 | - Present the individualized case management plan<br>- Identify and work to resolve barriers to accessing services<br>- Adjust case management plan to reflect current priorities |
| Week 4 | - Readminister social needs screener to identify new and ongoing social needs<br>- Identify and work to resolve barriers to accessing services<br>- Adjust case management plan to reflect current priorities |
| Week 6 | - Identify and work to resolve barriers to accessing services<br>- Adjust case management plan to reflect current priorities |
| Week 8 | - Readminister social needs screener to identify new and ongoing social needs<br>- Identify and work to resolve barriers to accessing services<br>- Adjust case management plan to reflect current priorities |
| Week 10 | - Identify and work to resolve barriers to accessing services<br>- Adjust case management plan to reflect current priorities |
| Week 12 | - Readminister social needs screener to identify new and ongoing social needs<br>- Identify and work to resolve barriers to accessing services<br>- Adjust case management plan to reflect current priorities<br>- Discuss with client long-term social needs and begin developing a long-term plan |
| Week 16 | - Readminister social needs screener to identify new and ongoing social needs<br>- Identify and work to resolve barriers to accessing services<br>- Adjust case management plan to reflect current priorities<br>- Review long-term plan and begin direct referrals to address long-term social needs |
| Week 20 | - Readminister social needs screener to identify new and ongoing social needs<br>- Identify and work to resolve barriers to accessing services<br>- Adjust case management plan to reflect immediate and long-term priorities |
| Week 24 | - Final check-in on referrals and outcomes<br>- Provide exit plan and long-term recommendations<br>- Administer participant satisfaction survey |

**Note:** Between weeks, the case manager makes direct referrals to community-based organizations on the participants' behalf. A participant may contact a case manager at any point between weeks to request additional meetings as needed.

## Measurements

Upon enrollment, participants will complete a baseline survey administered in REDCap that collects information on demographics, socioeconomic status, behavioral health, and experiences with and attitudes toward violence. For this study, the survey instrument was designed in conjunction with the coordinating center and five other research programs via a harmonization data collection.^22^

An overview of the survey domain included in the survey is provided in Appendix F. In addition to the baseline survey, participants will complete a social needs assessment and a violence risk assessment screener. The social needs assessment is used to identify social needs and to aid in the development of personalized case management plans for individuals enrolled in the Houston-HVIP (see Appendix E). The violence risk assessment, which is adapted from the Violent Reinjury Risk Assessment Instrument (VRRAI),^23^ is used to evaluate participants’ likelihood of future violence involvement and informs case managers on which participants would benefit from referrals to violence interrupter programs (see Appendix E). The VRRAI, initially developed by the UCSF-Wraparound Program, is the only comprehensive risk assessment tool designed specifically for HVIPs.

To track changes over time, participants will also complete follow-up surveys every three months for one year (i.e., at 3-month, 6-month, 9-month, and 12-month follow-ups). These follow-up assessments include time-varying measures from the baseline survey, allowing for examining changes in key outcomes over the study period. The survey and all data collection instruments are offered in English and Spanish.

## Primary Outcome

### Firearm Violence

The primary outcome will be a composite event defined as the occurrence of a firearm violence event within the 12-month follow-up period, identified by any of three data sources to triangulate data and enhance event capture: (1) EMR records from all Level-1 trauma centers in Harris County, TX, will be evaluated to assess individuals who return to the hospital with a new GSW, (2) mortality records from the national death index will be searched to identify participants who died as a result of a GSW during the follow-up period, and (3) self-reported data from a survey instrument will be captured during each follow-up assessment to measure firearm violence using items from the Conflict Tactic Scales (CTS2).^24^ Before answering the CTS2 portion of the survey, respondents will receive a prompt asking them to “Think about some of the behaviors that happened to you during any fights, conflicts, arguments, or physical attacks with” (a) friends, neighbors, co-workers, or strangers, and (b) with a boyfriend/girlfriend, fiancé/fiancée, or husband/wife. Respondents are then asked, “In the past 3 MONTHS, including TODAY, how often did each of the following things happen:”

a. Friends, neighbors, co-workers, or strangers threatened me with a gun or used/fired a gun on ME: (a) never, (b) once, (c) twice, (d) 3-5 times, (e) 6-10 times, (f) 11-20 times, or (f) 20+ times.
b. MY PARTNER threatened me with a gun or used/fired a gun on ME: (a) never, (b) once, (c) twice, (d) 3-5 times, (e) 6-10 times, (f) 11-20 times, or (f) 20+ times.
c. I threatened them with a gun or used/fired a gun at THEM: (a) never, (b) once, (c) twice, (d) 3-5 times, (e) 6-10 times, (f) 11-20 times, or (f) 20+ times.
d. I threatened them with a gun or used/fired a gun at MY PARTNER: (a) never, (b) once, (c) twice, (d) 3-5 times, (e) 6-10 times, (f) 11-20 times, or (f) 20+ times.

## Secondary Outcomes

### Firearm and Non-Firearm Victimization

Firearm and non-firearm victimization will be measured using the same criteria as the primary outcome, but will focus on information on cases pertaining to (a) hospital readmission for both firearm and non-firearm violent injury stemming from interpersonal violence (i.e., stabbings; assaults), (b) death due to a firearm or non-firearm violent injury, and (c) self-report for violence from firearms as well as non-firearms. Items from the CTS-2 will use the same prompts above and will include the following additional questions.

a. MY PARTNER pushed, shoved, or slapped ME: (a) never, (b) once, (c) twice, (d) 3-5 times, (e) 6-10 times, (f) 11-20 times, or (f) 20+ times.
b. MY PARTNER kicked, punched, beat me up, or threatened/used a knife or something sharp on ME: (a) never, (b) once, (c) twice, (d) 3-5 times, (e) 6-10 times, (f) 11-20 times, or (f) 20+ times.
c. THEY pushed, shoved, or slapped ME: (a) never, (b) once, (c) twice, (d) 3-5 times, (e) 6-10 times, (f) 11-20 times, or (f) 20+ times.
d. THEY kicked, punched, beat me up, or threatened/used a knife or something sharp on ME: (a) never, (b) once, (c) twice, (d) 3-5 times, (e) 6-10 times, (f) 11-20 times, or (f) 20+ times.

### Attitudes toward firearm violence

Attitudes toward violence are measured via self-reported survey items listed below, all of which include response options (strongly agree, agree, disagree, strongly disagree):^25^

a. It is okay to shoot a person if that is what it takes to get something you want.
b. It is okay to use a gun to shoot someone who insults or hurts you (for example: beats you up).
c. Carrying a gun makes you feel powerful/potent.
d. Carrying a gun makes you safer.
e. It’s okay to carry a gun if you live in a rough neighborhood.
f. It is okay to use a gun to threaten someone who disrespects you.

### Post-Traumatic Stress Disorder Symptoms (PTSD)

Symptoms of PTSD are measured using items from the Post-Traumatic Stress Disorder Checklist Version 2 (PCL-2).^26^ Specifically, respondents were asked in the past month how much they were bothered by (responses not at all, a little bit, quite a bit, extremely):

a. Repeated, disturbing, and unwanted memories of the stressful experience?
b. Feeling very upset when something reminded you of a stressful experience?

### Aggression

Aggression is measured using self-reported items from the Copeland-Linder Scale^27^ (response options: strongly agree, agree, disagree, strongly disagree):

a. I believe that if someone hits you, you should hit them back.
b. I believe revenge is a good thing.
c. I believe that it is OK to hurt people if they hurt you first.

### Self-Rated Health

Self-rated health is measured using a single survey item asking respondents “In general, would you say your health is?” (excellent, very good, good, fair, poor)

## Participant timeline

In addition to the baseline assessment, study participants will provide follow-up data at four additional time points: 3 months, 6 months, 9 months, and 12 months post-enrollment (see Figure 1 for details of the participant timeline). Surveys are administered electronically through REDCap. Survey compensation is $50 for all surveys. Following the final 12-month survey, participants will also be asked to complete a program satisfaction survey (see Appendix G).^28^ Additionally, a subset of participants (*n* = 40) will be invited to participate in semi-structured follow-up interviews, each lasting approximately 60 minutes, to gain deeper insights into their experiences with the intervention.^29^ Participants will be compensated $50 for the follow-up interview.

## Sample Size/Statistical Power

The sample size calculation is based on a comparison of the primary outcome of firearm violence between two randomization groups (Enhanced Usual Care vs. Houston-HVIP). We will extract the event dates for events captured in the first two data sources (EMR and mortality records), while the CTS2 measured at 3, 6, 9, and 12 months following enrollment will capture whether the event occurs between the visits. The first occurrence captured in the three data sources will then be determined for the composite event., We will compare the occurrence of firearm violence between the two arms using a Weibull regression model for the interval- and right-censored data to account for the interval-censoring (due to the planned measurement schedule for CTS2) and right-censoring (due to potential loss to follow-up and administrative censoring at 12 months). A prior study among youth seeking assault-injury care at an emergency department observed a two-year rate of 59% for a similarly-defined firearm violence composite, where the reporting rates between 0-6, 6-12, 12-18, and 18-24 months were 37.5%, 25.8%, 20.5%, and 15.2% respectively.^30^ This pattern suggests a potential decreasing hazard over time that can be well accounted for by a Weibull distribution, which flexibly allows for increasing, decreasing, or constant hazard shapes. The null hypothesis of no intervention effect corresponds to a regression coefficient of zero for the study group variable or equivalently a hazard ratio (HR) of 1.

We conducted statistical simulations based on a Weibull distribution with shape and scale parameters chosen according to rates in the prior study, such that the twelve-month probability is 42% in the SOC arm. To estimate the power, we generated 10,000 simulated data from this distribution, imposed interval censoring according to the planned follow-up, added right censoring from potential attrition (patient lost to follow-up), and applied the Weibull regression model with a two-tailed Wald test. Under a sample size of 274 (137 in each arm) and allowing up to 35% attrition during the follow-up, we have 80% power to detect a HR of 0.50 with a two-sided alpha of 0.05, corresponding to a reduction from 42% to 24% in twelve-month probability. The planned adjustment of important risk factors in the Weibull regression model will offer enhanced power. The planned sample size will also provide adequate power for the secondary outcomes. For example, for a continuous outcome, a complete sample size of 200 (100+100) allows us to detect a small to moderate between-group difference, Cohen’s d=0.40, with 80% power and a two-sided alpha=0.05 in a two-sample t-test.

## Consent

All participants will provide written informed consent and authorizations before randomization, in accordance with UTHealth institutional review board requirements. For individuals under 18, parental/legal guardian consent will be required, along with the minor’s assent, prior to randomization.

## Data Management

Data for this study will be collected using REDCap, with access restricted to the data manager, program manager, and case managers, all of whom will remain unblinded to study conditions. This limited access is intentional to minimize data entry errors, reduce the number of individuals responsible for data management, and maintain the integrity of the study by ensuring that personnel blinded to randomization conditions cannot access raw data files. To protect participant confidentiality, all data within the REDCap database will be de-identified. Through the data harmonization process in conjunction with the University of Michigan coordinating center for the CFVP Network, data will be stored on the Inter-University Consortium for Political and Social Research (ICPSR) at the end of this study and be made available to other researchers.

## Statistical Methods

The primary analysis will follow the intention-to-treat principle. A Weibull regression model for interval- and right-censored data will be used to compare the occurrence of firearm violence between the two study groups. A two-sided alpha of 0.05 will be used to determine statistical significance. Since prespecified adjustment of important baseline covariates can provide enhanced power, the regression model will be adjusted for race/ethnicity, biological sex, and age. In addition, we plan to balance-cross case managers across the two arms to protect against potential agent effects, if any, and will adjust the case manager as a fixed factor covariate in our analytic model.^31^

Next, to assess the potential heterogeneity of the intervention effect, we will examine interactions between the randomization group and important baseline risk factors (e.g., race/ethnicity) in additional secondary analyses. The corresponding interaction terms will be added individually to the regression model to examine potential effect modification. We will also carry out a per-protocol analysis as a sensitivity analysis.

The secondary outcomes that are measured longitudinally (e.g., PSTD) will be analyzed using appropriate longitudinal models, such as the (generalized) linear mixed model with a suitable distribution and link function, including group, time (categorical), group-by-time interaction, as well as subject-specific random effects to account for the repeated measures. We will first conduct a contrast to test the intervention effect across the post-baseline time points. Group-specific means and between-group differences will also be estimated at each time point with 95% confidence intervals.

## Ethical Considerations

All study procedures have been approved by the UTHealth Houston Committee for the Protection of Human Subjects (HSC-MS-23-0904) and the NIH. The study is registered on ClinicalTrials.gov (ID: NCT06263647). The study is overseen by a data monitoring and safety board at UTHealth Houston. Any adverse or serious adverse events will be reported to the study principal investigators, data monitoring and safety board, and institutional review board within 24 hours.

## Contingency Plans

In the event that participant recruitment falls short of the target enrollment of 274 individuals, we have developed a set of proposed contingency plans in consultation with the University of Michigan Coordinating Center and the Houston-HVIP Community Advisory Board. These plans are precautionary in nature and do not represent proposed changes to the UH3 phase at this time. Implementation of any contingency strategies will occur only with NIH approval and only if recruitment challenges arise under the current eligibility criteria.

1. **Expanding the Eligible Age Range:** We may consider increasing the upper age limit for participant eligibility from 35 to 40 years. Hospital data from 2024 suggest that this expansion would add approximately 89 additional eligible participants over an 18-month recruitment period.
2. **Broadening the Geographic Catchment Area:** Current eligibility is limited to residents of Harris County. TX. If necessary, we may expand eligibility to include individuals residing within a 40-mile radius of MHH–TMC, encompassing surrounding counties and municipalities. Based on 2024 data, this modification could add approximately 53 additional eligible participants aged 16–35.
3. **Extending the Recruitment Window:** The original study design specifies an 18-month recruitment period. If enrollment lags, we are prepared to extend this period up to 24 months. This adjustment would reduce the required average monthly enrollment from approximately 15 to 11 participants.
4. **Expanding Eligible Injury Types:** The current inclusion criteria are limited to individuals presenting with GSWs. If needed, we may expand eligibility to include patients with other penetrating violent injuries, such as stab wounds. Hospital data from 2024 indicate that this expansion would add approximately 105 eligible participants over an 18-month period.

## Discussion

### Strengths

This study design addresses several limitations identified in prior reviews of HVIPs.^13^ First, by implementing a two-arm RCT, this study may contribute to the HVIP literature by evaluating the effectiveness of intensive case management services compared to a control condition of usual care with a causal inference design. Second, many HVIP studies are limited by small sample sizes and insufficient statistical power, making it difficult to assess program effectiveness. In contrast, this study aims to recruit approximately 274 participants, providing adequate statistical power and positioning it as one of the largest HVIP RCTs conducted to date.^13^ Third, while most HVIP evaluations focus on reducing violent reinjury, few examine broader outcomes such as attitudes toward aggression, attitudes toward violence, as well as mental and overall health. Fourth, we employ a one-year follow-up period with data collection at 3, 6, 9, and 12 months, extending beyond the six-month post-injury follow-up period that is common in many prior HVIP studies.^32^ Fifth, our approach to measuring violent reinjury is multifaceted, leveraging (a) hospital readmission data covering both adult Level 1 trauma centers in the Greater Houston area, (b) comprehensive national death record searches, and (c) self-reported information using a validated instrument. Sixth, the intervention arm has also been designed to be client-centered to ensure that patients’ reported needs are considered and consistently updated so that social needs service referrals and connections address those underlying needs.^33^ Finally, responding to calls from scholars, this study includes a qualitative assessment to examine the impact of the HVIP program on participant experiences and outcomes.^29^

### Limitations

Despite the strengths of this study, there are limitations of the current study that can potentially be addressed by future HVIPs. First, the study focuses on GSW patients, limiting the generalizability of findings to individuals with other types of violent injuries, such as stabbings or assaults. Second, enrollment is restricted to individuals aged 16–35, a range selected based on hospital EMR data indicating the highest incidence of gunshot wounds within this age group.

The decision to enroll minors aged 16–17 was primarily because they are treated in the adult trauma unit while enrolling patients under 16 would require coordination with the affiliated children’s hospital trauma unit. Consequently, the findings may not be applicable to GSW victims outside of the targeted age range. Third, study participation is limited to individuals who are fluent in English or Spanish. While this criterion encompasses the vast majority of GSW patients presenting to the hospital, it restricts the ability to assess HVIP effectiveness among those who speak other languages. Additionally, the study excludes individuals with self-inflicted or unintentional gunshot wounds, preventing conclusions about the role of HVIPs in reducing the recurrence of these types of injuries. Finally, the study is conducted at a large, urban, Level 1 trauma center, which provides a high volume of GSW patients for recruitment. However, this setting may limit the generalizability of findings to other geographic regions or hospital types.

### Dissemination

Findings will be disseminated through peer-reviewed journal publications, meetings with local hospital administrators, and presentations at local, regional, and national conferences.

## CONCLUSION

Firearm violence imposes substantial social and economic burdens on society. In response, HVIPs have been increasingly implemented in cities across the US to support victims of violence by addressing their needs before discharge. However, despite the growing body of research on HVIPs, their effectiveness remains inconsistent, and there is a lack of large-scale RCTs assessing their impact on violent reinjury.^13^ To fill this gap, we anticipate conducting an RCT at a major Level 1 trauma center in the fourth-largest city in the United States. Findings from this study will provide critical insights to inform the design and implementation of future HVIP programs.

## Data Availability

No data are reported yet. Data will be made available in ICPSR.

### Appendix A: Distribution of GSW Patients by Age in 2024 (*N* = 685)

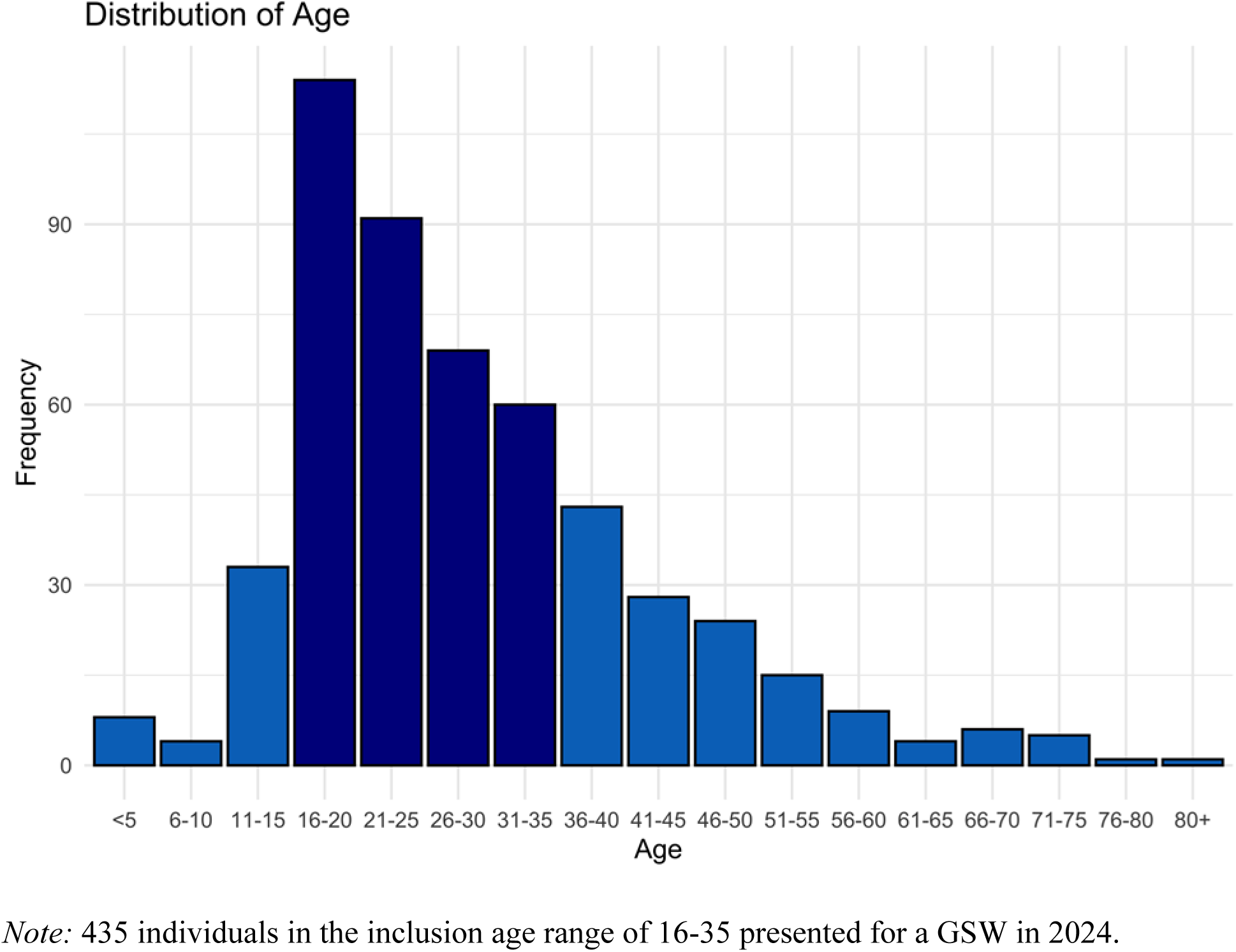

### Appendix B: Rapport Building Script

*This is a **<u>GUIDE</u>** that the case manager can use during the first interaction with the patient when the patient is medically stable. Recruitment and retention success highly depends on the initial contact and the relationship we establish with the patient/participant. It is important that case managers and study team members build trust with the patients/participants by making eye contact when speaking with them and letting them know that you’re listening to them. It is important to connect with the patient by asking questions and support them by responding to their needs and requests*.

### Introduction

Hi my name is (*case manager’s name*), and I am a case manager at UTHealth. Is it OK if I call you (check with clinical staff/EMR))? I am working on a program to help individuals who have experienced gun injuries and will try to connect you to resources to help you. I’m also here to offer support and gather some information that will help us improve our services for patients in similar positions.

#### Building rapport

*Express Empathy:* " How are you feeling today, both physically and emotionally?"

*Show Genuine Concern:* "It’s important to us that you feel supported during your recovery. Is there anything specific you need right now or any immediate concerns you’d like to talk about?" Is there anyone you’d like us to contact right now?

If you don’t mind my asking, can you tell me about the events that led to your hospital visit?

*Listen Actively:* "How do you feel like you are/were treated when you were admitted to the hospital?

*Personal Connection:* “Do you have a place to stay right now? What is your work situation? If you’re comfortable, could you share a little about yourself, such as your interests or what you enjoy doing?"

Interaction with Medical Staff:

*Build Trust:* "How would you describe your experience with the medical staff? Were they understanding and professional in regard to your needs/care?"

*Ensure Clarity:* "Were you given clear and understandable information about your diagnosis and treatment plan?"

Concerns and Needs:

*Acknowledge Their Concerns:* "Were there any specific concerns or difficulties you encountered during your hospital visit that we haven’t discussed that you would like me to address? How can we address them better in the future?"

*Offer Reassurance:* "Our goal is to ensure you feel heard and supported. Did you feel that your needs were met during your time in the hospital?"

#### After the patient completes the survey/before the case manager leaves the participant

"Thank you so much for taking the time to share your experiences with me. Your feedback is crucial in helping us improve our services and support for other gunshot wound patients. If you have any further questions or need additional support, please don’t hesitate to reach out. I hope you have a speedy recovery.”

### Appendix C: Eligibility Screening Form

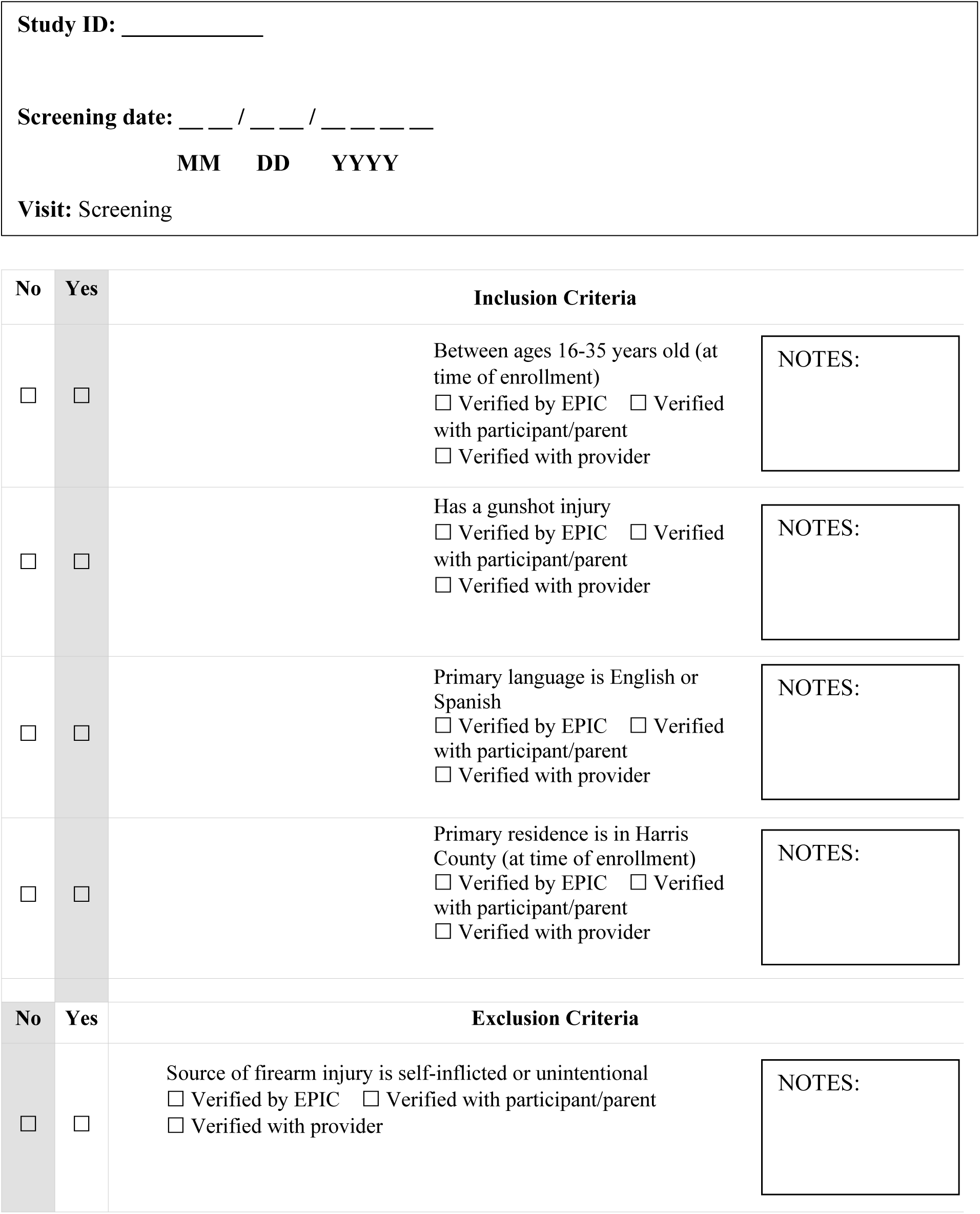
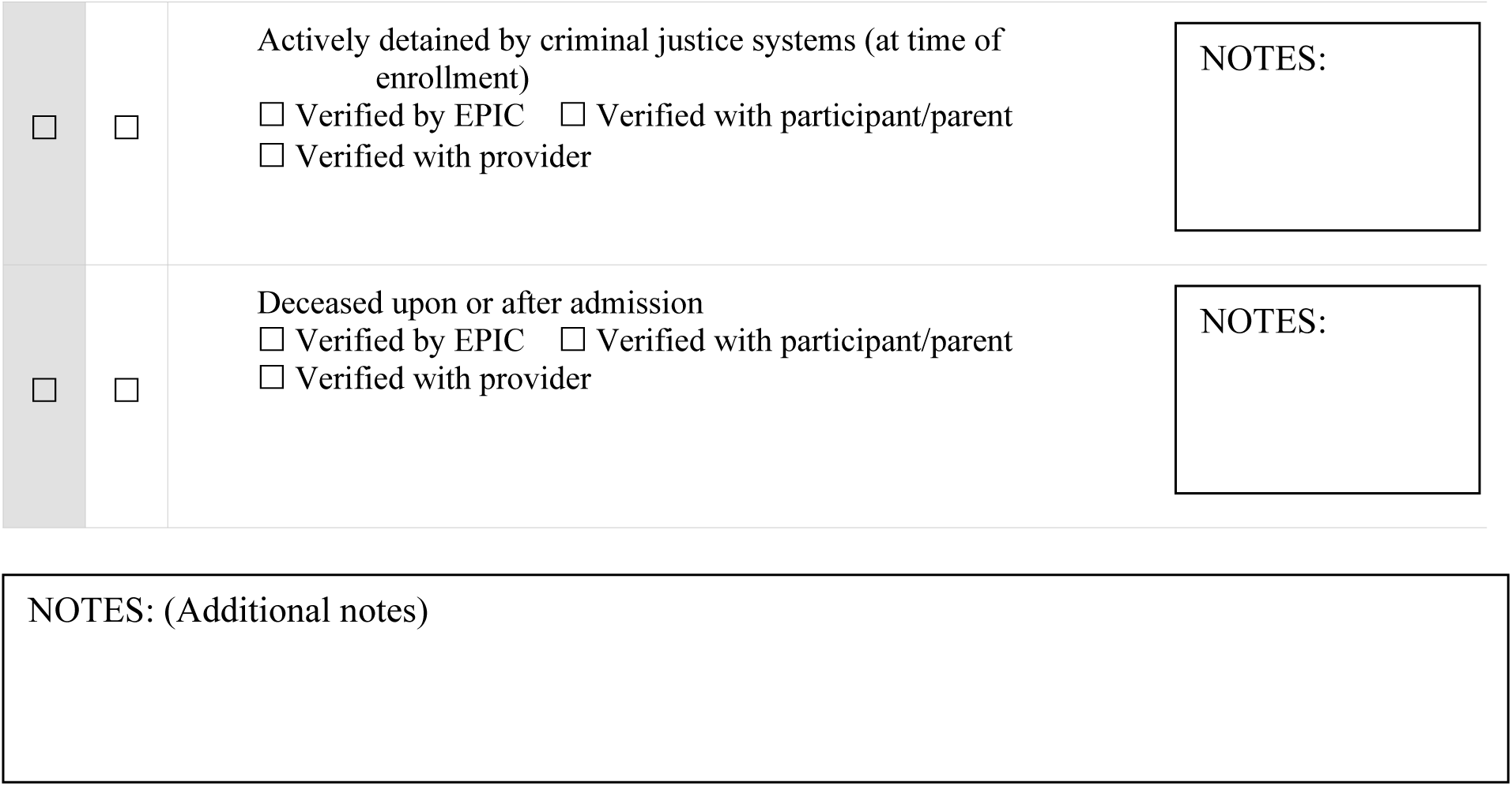

### Appendix D: Social Risk Screening

Please tell me if you are currently concerned about any of the following for yourself. The answers are YES/No.

|  |  |  |
| --- | --- | --- |
| 1. Running out of food before having enough money or food stamps to buy more. | If YES, would you like help with this? | ☐ Yes ☐ No ☐ Maybe |
| 2. Unstable housing, including eviction, foreclosure, homelessness, or staying with friends/family. | If YES, would you like help with this? | ☐ Yes ☐ No ☐ Maybe |
| 3. Problems paying bills, like electric, gas, water, or phone bills | If YES, would you like help with this? | ☐ Yes ☐ No ☐ Maybe |
| 4. Housing problems like mold, insects, rats, or mice | If YES, would you like help with this? | ☐ Yes ☐ No ☐ Maybe |
| 5. Difficulty finding a job | If YES, would you like help with this? | ☐ Yes ☐ No ☐ Maybe |
| 6. Do you interested in starting or continuing school or job training? or example, working toward a high school diploma, GED, or completing a training program | If YES, would you like help with this? | ☐ Yes ☐ No ☐ Maybe |
| 7. Difficulty quitting or cutting down on using too much alcohol or drugs | If YES, would you like help with this? | ☐ Yes ☐ No ☐ Maybe |
| 8. A disability interfering with the ability to work | If YES, would you like help with this? | ☐ Yes ☐ No ☐ Maybe |
| 9. Problems with a current or former job, like unpaid wages, workers comp, discrimination or harassment | If YES, would you like help with this? | ☐ Yes ☐ No ☐ Maybe |
| 10. Difficulty obtaining unemployment insurance | If YES, would you like help with this? | ☐ Yes ☐ No ☐ Maybe |
| 11. Getting cut off from or denied from programs that provide income support, like the Supplemental Nutrition Assistance Program (food stamps) or unemployment benefits. | If YES, would you like help with this? | ☐ Yes ☐ No ☐ Maybe |
| 12. Having no health insurance | If YES, would you like help with this? | ☐ Yes ☐ No ☐ Maybe |
| 13. Having no primary care provider | If YES, would you like help with this? | ☐ Yes ☐ No ☐ Maybe |
| 14. Receiving medical or pharmacy bills that you cannot afford | If YES, would you like help with this? | ☐ Yes ☐ No ☐ Maybe |
| 15. Difficulty finding childcare | If YES, would you like help with this? | ☐ Yes ☐ No ☐ Maybe |
| 16. Concerns about your mental health | If YES, would you like help with this? | ☐ Yes ☐ No ☐ Maybe |
| 17. Difficulty with transportation | If YES, would you like help with this? | ☐ Yes ☐ No ☐ Maybe |
| 18. Concerns about your safety | If YES, would you like help with this? | ☐ Yes ☐ No ☐ Maybe |

*Note:* The screening tool is adapted from the following studies:
Garg, A., Toy, S., Tripodis, Y., Silverstein, M., & Freeman, E. (2015). Addressing social determinants of health at well childcare visits: a cluster RCT. *Pediatrics*, 135(2), e296-e304.
Gottlieb, L. M., Adler, N. E., Wing, H., Velazquez, D., Keeton, V., Romero, A., Hernandez, M., Vera, A. M., Caceres, E. U., Arevalo, C., Herrera, P., Suarez, M. B., & Hessler, D. (2020). Effects of in-person assistance vs personalized written resources about social services on household social risks and child and caregiver health: a randomized clinical trial. *JAMA Network Open*, 3(3), e200701-e20070

### Appendix E: Risk Assessment

1. Do you currently feel that you or someone close to you is in immediate danger of being harmed due to an ongoing conflict?

☐ Yes
☐ No
2. Are you a member of a gang or do you regularly spend time with individuals who are associated with a gang?

☐ Yes
☐ No
3. Including the event that led you to the hospital how many times have you previously been (a) shot, or (b) stabbed?

**3.1** Number of shootings:
**3.2** Number of Stabbings:

### Personal incarceration

#### Family History of Incarceration Study (FamHIS) - Release

4. Have you ever spent time in a jail, prison, juvenile detention center or other correctional facility?

☐ Yes
☐ No
4.1 Was your incarceration related to a violent crime (e.g., assault, robbery, or other offenses involving physical harm or the threat of harm)?

☐ Yes
☐ No

These next questions ask about people you may have known who were killed by a gun. We know these are personal questions and all your answers are private.

If you are experiencing any distress related to this question, please call either the National Crisis Hotline at 988 or notify the Research Assistant (RA) and we will provide helpful resources.

5. How many of your friends or family members (for example, your Mom, Dad, Sister/Brother, Cousin) have ever been shot or killed with a gun?

☐ 0
☐ ….
☐ 25+

*Note:* Screening tool is adapted from: Kramer, E. J., Dodington, J., Hunt, A., Henderson, T., Nwabuo, A., Dicker, R., & Juillard, C. (2017). Violent reinjury risk assessment instrument (VRRAI) for hospital-based violence intervention programs. *Journal of Surgical Research*, *217*, 177-186.

### Appendix F: Houston-HVIP Baseline Survey Domains

| **Demographic Characteristics:**<br>Age<br>Education<br>School<br>Employment Status<br>Employment Hours<br>Employment Type<br>Household Income<br>Household size<br>Children in the home<br>Race<br>Sex<br>Sexual orientation<br>Relationship status | **Non-firearm Violence:**<br>Community violence exposure<br>Partner and Non-partner victimization<br>Partner and Non-partner aggression<br>Physical Fighting<br>Attitudes toward aggression and violence<br>Criminal justice consequences<br>Avoidance behaviors<br>Violence prevention self-efficacy | **Firearms and firearm violence:**<br>Firearm violence exposure<br>Firearm violence attitudes<br>Firearm ownership<br>Firearm carriage<br>Firearm carriage reasons<br>Risky firearm behaviors<br>Risky Firearm Discharge Behaviors<br>SaFETy score |
| --- | --- | --- |
| **Social Risk Factors** | **Adverse Childhood Experiences** |  |
| **Personal incarceration** | **Social media** |  |
| **Mental Health:**<br>Depression<br>Anxiety<br>Loneliness<br>PTSD | **Risk Assessment** |  |
| **Substance Use:**<br>Alcohol use<br>Drug use |  |  |
| **Physical Health:**<br>General Health<br>Sleep medication<br>Daytime dysfunction |  |  |
| **Resilience** |  |  |

### Appendix G: Patient Satisfaction Survey

| Item | Response Options |
| --- | --- |
| Using any number from 0 to 10, what number would you use to rate the services you received from (case manager name) while in the program? | 0 (Worst)–10 (Best) |
| To what extent has our program met your needs? | All my needs were met, Most of my needs were met, A few of my needs were met, None of my needs were met |
| How happy are you with the amount of help you received? | Very happy, Mostly happy, Neither happy nor unhappy, Quite unhappy |
| Did the program help you to feel better? | It helped me a great deal, It helped somewhat, It did not help, It made me feel worse |
| Overall, how happy were you with the services you received? | Very happy, Mostly happy, Neither happy nor unhappy, Quite unhappy |
| If you were to experience violence again, would you choose to come back to our program? | Definitely, Maybe, Probably not, Definitely not |
| How often did the program staff listen carefully to you? | Always, Usually, Sometimes, Never |
| Program staff respect parts of your culture or identity that are important to you. | Always, Usually, Sometimes, Never |
| Because of the program, you know more about community resources that are available to you. | Definitely, Maybe, Probably not, Definitely not |
| Because of the program, you feel better prepared to access community resources, such as legal, medical, educational, or mental health services. | Definitely, Maybe, Probably not, Definitely not |
| You are prepared to apply the skills and tools you learned during the program. | Definitely, Maybe, Probably not, Definitely not |
| Is there anything else you would like to share about (program name), such as what was helpful or unhelpful or ways that (program name) could be improved? | Free text |

Note: Survey adapted from: Myers, R. K., Kapa, H. M., Garcia, S. M., Vega, L., & Fein, J. A. (2025). Development of a Brief Client Satisfaction and Quality Improvement Tool for Hospital-Based Violence Intervention Programs: Opportunities for Enhancing Client Perspectives. *Journal of Patient Experience*, 12, 23743735251314622.

